# Dorsal Subthalamic Deep Brain Stimulation Improves Pain in Parkinson’s Disease

**DOI:** 10.1101/2022.05.30.22275774

**Authors:** Asra Askari, Jordan Lam, Brandon J. Zhu, Charles Lu, Kelvin L. Chou, Kara J. Wyant, Parag G. Patil

## Abstract

**Introduction:** Inconsistent effects of subthalamic deep brain stimulation (STN DBS) on pain, a common non-motor symptom of Parkinson’s disease (PD), may be due to variations in active contact location relative to a pain-reducing locus of stimulation.

**Objective:** To distinguish the loci of maximal effect for pain and motor improvement in the STN region.

**Methods:** We measured Movement Disorder Society Unified PD Rating Scale (MDS-UPDRS) Part I pain score (item-9), and MDS-UPDRS Part III motor score, preoperatively and 6-12 months after STN DBS. An ordinary least-squares regression model was used to examine active contact location as a predictor of follow-up pain score while controlling for baseline pain, age, dopaminergic medication, and motor improvement. An atlas-independent electric field model was applied to distinguish sites of maximally effective stimulation for pain and motor improvement.

**Results:** In 74 PD patients, mean pain score significantly improved after STN DBS (*p* = 0.01). In a regression model, more dorsal active contact location was the only significant predictor of pain improvement (*R*^2^ = 0.17, *p =* 0.03). The stimulation locus for maximal pain improvement was lateral, anterior, and dorsal to that for maximal motor improvement.

**Conclusions:** More dorsal STN DBS improves pain. Stimulation of the zona incerta, a region known to modulate pain in humans, may explain this observation.

## Introduction

Pain is a common and distressing non-motor symptom of Parkinson’s disease (PD), affecting up to 85% of patients and impacting quality of life [1]. Subthalamic nucleus deep brain stimulation (STN DBS) improves global pain scores from 28% to 84% [2]. However, the structure-function relationships underlying this finding remain unclear. Several studies correlate motor improvement and pain relief, while other studies find no relationship between motor and pain improvement after DBS [3-5].

Recent electrophysiologic studies have suggested distinct roles of STN DBS on pain and motor improvement [2, 6-8]. Connections between STN subregions and brain areas involved in pain processing [2, 9-12] and the proximity of motor-optimal DBS stimulation loci to the zona incerta (ZI), a potential target for pain relief [13, 14], motivated us to investigate the role of active contact location on pain in PD patients undergoing STN DBS.

In this study, we evaluated the impact of active DBS contact location on changes in the MDS-UPDRS Part I pain score following STN DBS. We hypothesized, based on earlier work [[14]], that stimulation of the dorsal STN/zona incerta (ZI) could improve pain scores.

## Methods

### Participants

This retrospective observational study included patients with idiopathic PD who underwent STN DBS at the University of Michigan between 2009 and 2019 and completed the MDS-UPDRS Part I questionnaire. Written informed consent was obtained from all subjects. The study was approved by the Institutional Review Board.

### Surgical Procedure

Several weeks before surgery, patients underwent a 3T MRI to visualize the STN, using a validated, high-resolution protocol [15]. On the day of surgery, patients were fitted with a Leksell stereotactic frame (Elekta Instruments AB, Stockholm, Sweden) and underwent a 1.5T MRI. The 3T MRI and 1.5T MRI were co-registered using a mutual-information algorithm (Analyze 9.0; AnalyzeDirect, Inc, Overland Park, Kansas). The MR-visualized STN was then targeted, and localization was finalized with intraoperative microelectrode recording. During surgery, a movement disorders neurologist evaluated each patient intraoperatively for symptom improvement and side effect thresholds to optimize DBS lead placement. At the time of pulse generator implantation, 2-4 weeks after lead placement, a high-resolution CT scan (CT750 HD, GE Healthcare, Chicago, Illinois; 64-slice, 140 kV, 450 mA, 0.5 × 0.5 × 0.6 mm) was obtained to visualize electrode contacts after brain shift and pneumocephalus had resolved. DBS programming commenced 4-6 weeks after initial lead placement. Our detailed surgical protocol is described in a previous publication [16].

### Active Contact Localization

Co-registered CT and MR images were oriented in Talairach space. The STN midpoint was defined as the point halfway between the STN oral and caudal poles [17], which were identified on coronal MRI. Coordinates of the active contacts were then determined and recorded relative to these MR-visualized STN midpoints. Lateral (X), anterior (Y), and dorsal (Z) directions relative to the STN midpoint were defined as positive.

### Determination of Loci of Maximal Effect

To determine the loci of maximal effect for motor improvement and pain relief, a weighted score was generated at each coordinate around the STN using the location of the active contact, the motor improvement score or the pain relief score, and the probability that the active contact would activate a neuron at the coordinate. An overall stimulation-weighted improvement score was assigned to each point in the STN region for each condition by summing across all patients. The simplex algorithm (MATLAB, MathWorks®, Natick, MA) was used to identify the locus associated with maximal motor or pain improvement. Spatial distributions (relative to STN midpoint in millimeters) and confidence intervals (CI) for maximal effect locations were calculated using the bootstrap technique. See Conrad et al. [16, 18] for greater detail.

### Clinical Assessments

Demographic information and levodopa-equivalent dose (LED) were collected. Clinical assessments included the MDS-UPDRS Part I pain score (item-9) and the MDS-UPDRS Part III (motor examination), which was measured before surgery in the OFF-medication condition (baseline) and 6-12 months later in the OFF-medication/ON-stimulation state. To complete the MDS-UPDRS Part I-9, patients were asked whether they have had any uncomfortable feelings in their body including pain, aches, tingling, or cramps over the past week, and then they scored from 0 (no uncomfortable feeling) to 4 (severe, that stoped them to do their daily activity of life) [19].

### Statistical Approach

An ordinary least squares regression model was used to examine the impact of active contact electrode location in each axis on pain score at follow-up while controlling for gender, age, LED change from baseline to follow-up, MDS-UPDRS-III percent change improvement, and pain score at baseline. Student’s paired t-tests were used to determine if the post-DBS assessments significantly differed from baseline assessments. All analyses were 2-sided with a significance level of 0.05.

## Results

### Participants

Our sample population included 74 patients with idiopathic PD who underwent STN DBS (72 with bilateral implants, 2 with unilateral). Twenty-two (29.7%) patients were female. The mean age was 64.2 ± 7.7 years. There was a small but significant (*p* = 0.01) improvement from baseline pain (1.87 ± 1.16) to follow-up pain score (1.41 ± 1.07).

### Relationship Between Pain Relief and Dorsal Active Contact Location

Regression analyses were conducted separately for the left and right hemispheres with lead location predictors separated by axis, resulting in a total of 6 analyses ([right or left hemisphere] x [X, Y, or Z axis]). The results of the ordinary least square regression model revealed that age, LED change from baseline to follow-up, gender, and pain score at baseline were not significant predictors of pain score at follow-up in any model (*p* > 0.05).

Active contact location in the Z-axis of the left hemisphere significantly predicted pain improvement (β = −0.2, *R*^2^ = 0.17, *p =* 0.03), with dorsal location providing greater pain relief. By contrast, active contact location relative to the STN midpoint in the X and Y axes in both hemispheres, and the Z-axis in the right hemisphere did not predict pain improvement (*p* > 0.05). In addition, the correlation coefficient between the pain follow-up score and left electrode active contact location in the Z-axis was highly significant (*R* = −0.37, *p =* 0.001; Figure 1).

**Figure 1.**
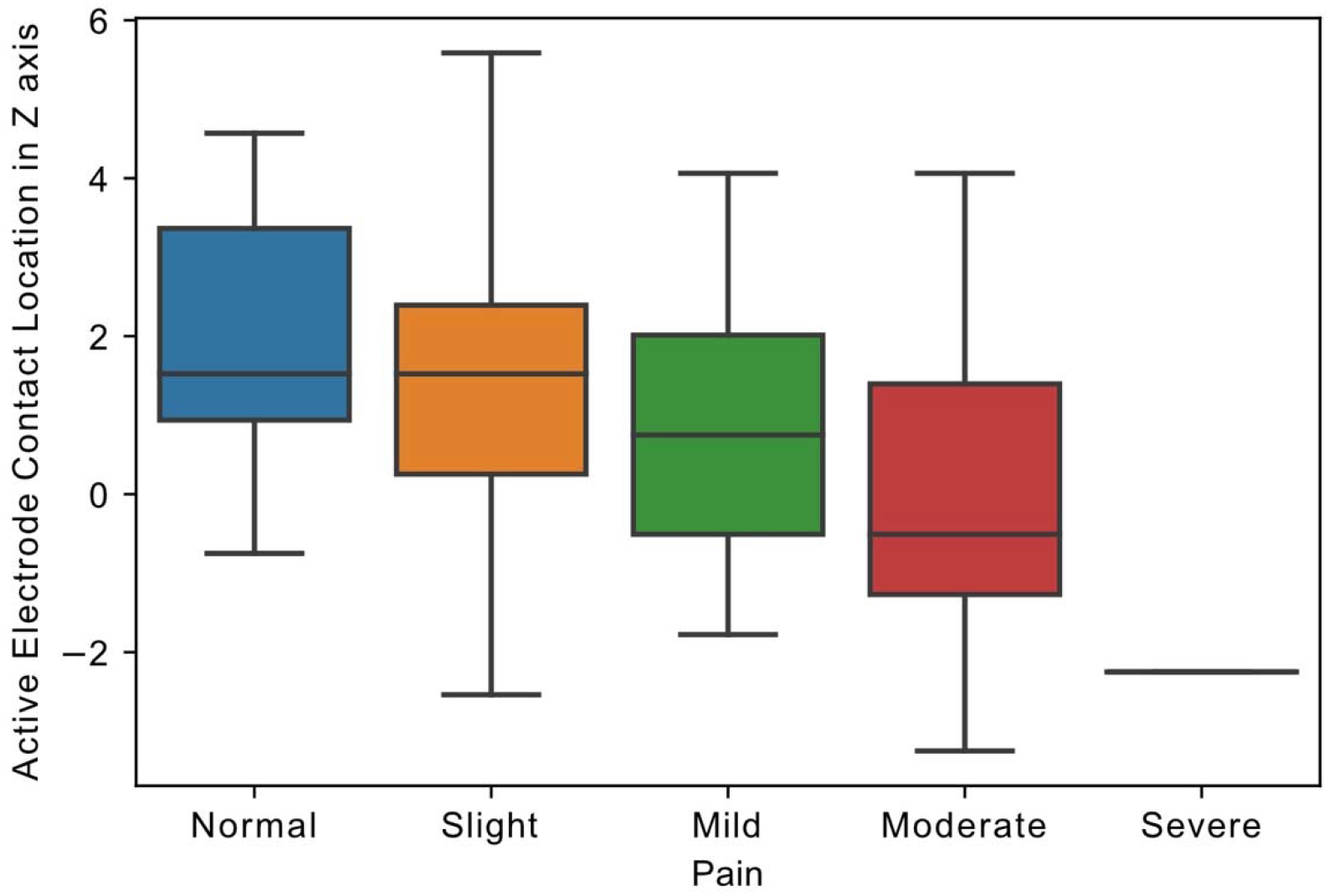
Relationship of Active Electrode Location to Pain. Boxplot, showing the distribution of follow-up pain scores and the distance of electrode active contact from STN midpoint in Z-axis in the left hemisphere (in millimeters). Pain scores in MDS-UPDRS Part I ranged from 0 to 4, representing 0: Normal, 1: Slight, 2: Mild, 3: Moderate, and 4: Severe pain.

### Loci of Maximal Motor and Pain Improvement

The locus of maximal effect for motor improvement was located medial (−0.22 mm, 95% CI [0.50, 0.04]), posterior (−0.70, [-1.10, -0.31]), and dorsal (0.98, [0.63, 1.34]) to the STN midpoint, while the locus of maximal effect for pain relief was lateral (0.15, [-0.75, 1.18]), anterior (0.57, [-0.85, 2.03]), and dorsal (2.26, [1.23, 4.09]) to the STN midpoint (Fig. 2). All coordinates are calculated in millimeters, with 95% confidence intervals, relative to the MR-visualized STN midpoint.

**Figure 2.**
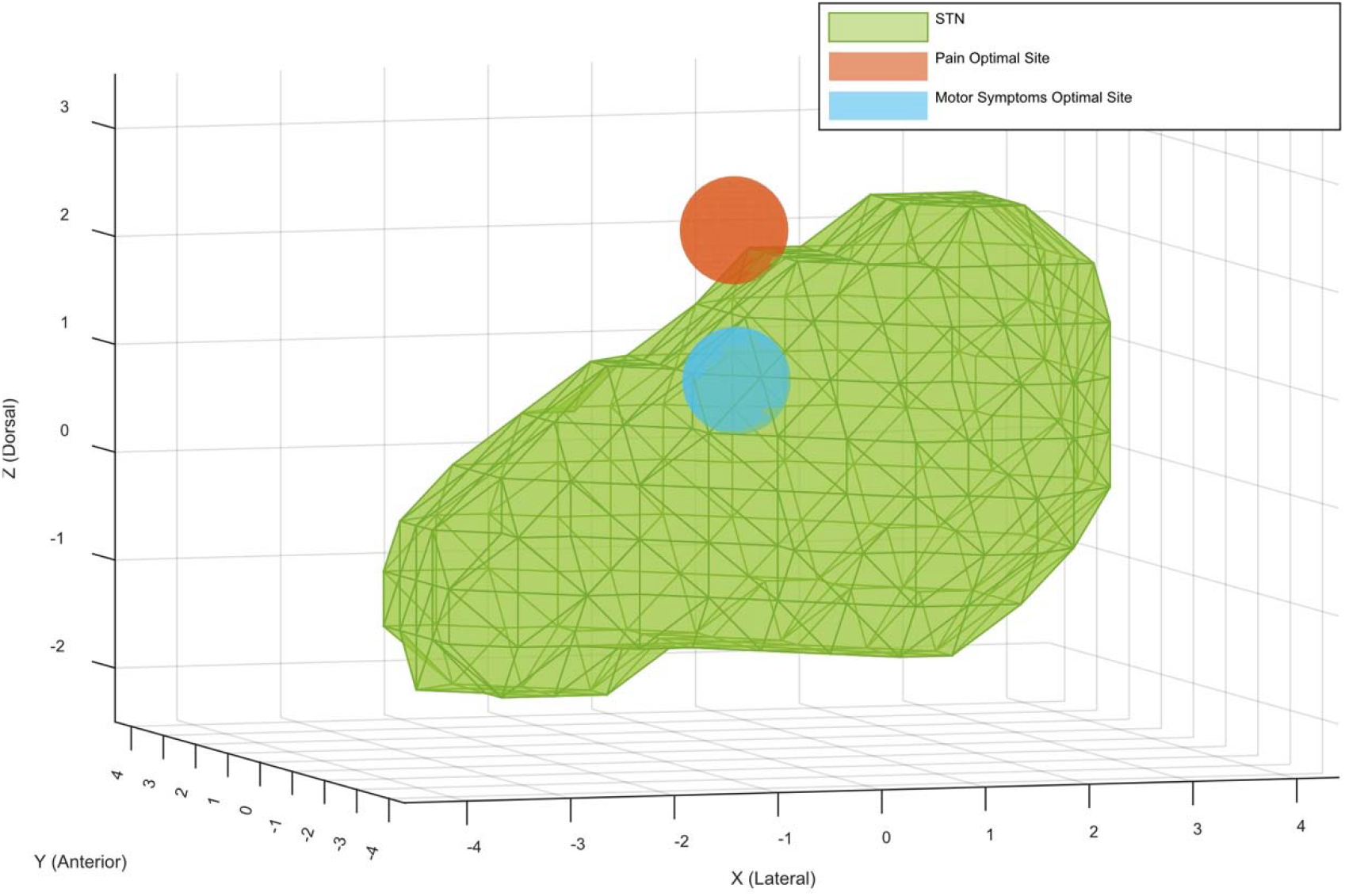
Locus of Optimal Pain and Motor Improvements in STN DBS. Mean coordinates for computational model-derived optimal sites of stimulation for pain and motor symptoms superimposed with the STN. The coordinates for the optimal site for pain are (X = 0.15 mm, Y = 0.57 mm, Z = 2.26 mm), and the coordinates for the optimal site for motor symptoms are (X = - 0.22 mm, Y = -0.70 mm, Z = 0.98 mm) relative to the STN midpoint. All axis coordinates are in millimeters, and lateral, anterior, and dorsal are defined to be positive.

## Discussion

We find that STN DBS significantly but modestly improves pain in PD patients. Dorsal active contact location is the only significant predictor for improvement in pain, independent of age, gender, motor improvement, or LED changes. Moving from the ventral to the dorsal STN subregion is associated with decreasing pain; with the site of maximal pain relief lying lateral, anterior, and dorsal to the site of maximal motor improvement. There was no association between right-sided active contact location and pain score at follow-up. The optimal motor response was observed in left-sided and bilateral stimulation and not in right-sided stimulation [20]. Likewise, the adverse effect of STN DBS is more significant for left-sided stimulation [21]. Zhang and colleagues showed the laterality for nociceptive perception [22]. However, additional study is needed to investigate pain perception laterality and the differential effects of left- and right-sided stimulation.

There are several detailed rating scales evaluating pain intensity and frequency in PD, thus recently the King’s scale has been introduced as one of the most accurate [23]. However, to our knowledge, there were no studies assessing the association between the STN subregions and a detailed pain rating scale in a large cohort. However, MDS-UPDRS part I-9 is not a detailed questionnaire; it has been validated [24] and showed that was highly associated with quality of life [25].

Pain in PD is multifactorial, with evidence to support a nondopaminergic responsive central component [2, 26, 27]. The role of ZI, the gray matter band located dorsal to STN, in central pain processing has been previously described [13, 28]. Notably, the ventral subregion of ZI, enriched with GABAergic cells and connected to the spinothalamic tract and sensory thalamus, is an area involved in pain processing and modulation [29]. We recently found that low-frequency, 20 Hz stimulation of zona incerta (ZI), modulates heat pain in humans [14]. This finding supports the possibility of pain processing in ZI, specifically in PD patients. With the proximity of the dorsal STN subregion to ZI, we hypothesized that the spread of high-frequency current to the ZI ventral subregion could explain the positive effect of STN DBS on pain. The effect of low in comparison with high-frequency ZI stimulation on pain in PD needs to be addressed.

Neurostimulation of the dorsal STN subregion may also directly modulate pain processing. The functional connections between STN and pain processing regions in cortex, pedunculopontine nucleus, and parabrachial nucleus may play a role in this phenomenon [2]. Additionally, STN local field potentials have been shown to respond to pain stimuli [30]. However, the specific territories of STN involved in pain processing are poorly described. Future electrophysiologic studies could investigate the potential therapeutic role of the dorsal STN stimulation on pain.

Our study has several limitations that are needed to address. We only used one item of the MDS-UPDRS-I to evaluate pain, so we cannot say if DBS improves certain types of pain more than others. Future prospective studies could investigate the role of active contact location on improving different categories of pain by applying a detailed pain battery such as the King scale [31]. We applied an electric field model to localize the maximal effect site for pain improvement. However, directly simulating the volume of tissue activated around active contacts could be a more precise method to determine the optimal site for pain improvement. Although the maximal effect site was located at the uppermost dorsal subregion, its overlap with ventral ZI was undetermined. Future electrophysiologic studies and functional imaging could address this uncertainty.

To conclude, Stimulation of the STN dorsal subregion was associated with the improvement of MDS-UPDRS part I-9, a comprehensive rating scale measuring the impact of pain on quality of life. This may be related to stimulation in the vicinity of the ventral ZI, or direct pain-ameliorating effects on the dorsal STN. This finding motivates future studies to assess the effect of electrode active contact location on pain and its subdomains with a more detailed pain scale.

## Data Availability

All data produced in the present study are available upon reasonable request to the authors and IRB approval

## Author contributions

The study was conceived and designed by all authors. Data were acquired by AA, analyzed by AA and BJZ, and interpreted by all authors. The manuscript was written by AA. All authors reviewed the manuscript.

## Datasets availability

The raw data supporting the conclusions of this article will be made available by the authors, without undue reservation.

## Declarations

### Ethics approval

University of Michigan Institutional Review Board, approval HUM00021058.

### Consent to participate

Written informed consent was obtained from all subjects.

